# Serum brain-derived tau distinguishes acute neurotrauma diagnoses

**DOI:** 10.64898/2026.11.08.26365143

**Authors:** Ruyu Shi, Sarah E. Svirsky, Wasiu G. Balogun, Xuemei Zeng, John D. Fedak, Michel Nafash, Shayna T. Brodman, David O. Okonkwo, Ava M. Puccio, Thomas K. Karikari

## Abstract

Traumatic brain injury (TBI) and spinal cord injury (SCI) are two common forms of neurotrauma that can present with overlapping clinical features, complicating early diagnosis and management particularly in acute and mixed-trauma settings.^1–3^ Accurate and rapid biological differentiation remains an unmet need.^4^ We hypothesized that brain-derived tau (BD-tau), an ultrasensitive blood-based biomarker preferentially selective for tau protein originating from the brain, would distinguish TBI from SCI. In a multi-cohort assessment of 342 patients admitted to the neuro-intensive care unit (285 with TBI, 57 with SCI), and 22 controls, baseline serum BD-tau concentrations were markedly elevated in TBI compared with SCI, discriminating between the two populations with high accuracy (area under the curve [AUC]=0.91, 95%CI 0.87-0.96). In paired serum-cerebrospinal fluid (CSF) samples, serum BD-tau significantly outperformed CSF BD-tau for injury-type discrimination (AUC=0.95, 95%CI 0.89-0.99 vs. 0.80, 95%CI 0.71-0.89). Baseline serum BD-tau also showed superior discrimination of TBI from SCI compared with established blood biomarkers of neurotrauma, including neurofilament light chain, glial fibrillary acidic protein, and ubiquitin C-terminal hydrolase L1 (AUC=0.82, 95%CI 0.76-0.88; vs. 0.53, 95%CI 0.43-0.63; 0.67, 95%CI 0.58-0.76; 0.71, 95%CI 0.64-0.79; all DeLong p<0.001). Longitudinally, serum BD-tau remained persistently elevated in TBI, but not SCI, for up to 144 hours post-injury, with distinct temporal trajectories between injury types. Together, these findings indicate that serum BD-tau enables rapid and minimally invasive differentiation of TBI from SCI and may support early clinical decision-making, triage, and enrollment into injury-specific therapeutic trials, highlighting the potential of circulating biomarkers to improve precision diagnostics in acute neurotrauma care.

## Main

Traumatic brain injury (TBI) and spinal cord injury (SCI) are the first and second leading causes of disability and morbidity in the United States.^5–7^ The gold standard methods to assess injury severity and classification are the Glasgow Coma Scale (GCS) score and Computed Tomography (CT) neuroimaging for TBI and the American Spinal Injury Association (ASIA) Impairment scale for SCI ^8–10^. However, these baseline examinations have varied accuracies in the acute care setting or can be unobtainable due to the unstable condition of the patient.^11,12^ This is further complicated by substantial differences in clinical management, prognostication, and treatment indications differ substantially between TBI and SCI.^13–16^ Therefore, there is a crucial need for rapid, reliable, and accurate tools to support the confirmation and differential diagnosis of TBI and SCI.

Blood-based biomarkers show promise for improving diagnosis, longitudinal monitoring and prognosis of patients with TBI and SCI.^17,18^ Several blood biomarkers that reflect the complex and dynamic acute pathophysiology and temporal trends of TBI and SCI is already available.^19,20^ For example, plasma/serum glial fibrillary acidic protein (GFAP) and ubiquitin C-terminal Hydrolase L1 (UCH-L1) increase immediately post-injury in response to astrocytic and neuronal cell injury/death and are cleared by the US Food and Drugs Administration to aid in the diagnosis of mild TBI and for determining need of a CT scan among adults.^21,22^ Additionally, plasma/serum neurofilament light chain (NfL) and total tau (t-tau) show promise as blood biomarkers of axonal injury.^23^ However, these blood biomarkers are not ideal. The most notable limitation is their inability to differentiate between common acute neurotrauma disorders such as TBI and SCI, as they are similarly elevated in both conditions.^24–26^ Moreover, plasma/serum GFAP, UCHL-1 and t-tau levels increase acutely, however contributions from systemic sources reduce confidence in diagnostic utility and specificity for central nervous system (CNS) disorders.^27,28^ Plasma NfL levels peak in the sub-acute period, limiting its use as an acute marker for rapid triage.^29^ Additionally, the levels of these proteins are affected by co-morbid conditions and normal aging.^30–32^

We recently described brain-derived tau (BD-tau), a novel blood-based biomarker that selectively recognizes tau protein isoforms released into blood circulation predominantly from the brain.^33,34^ This biomarker assay was designed to recognize the tau protein isoforms abundantly expressed in the adult human brain^34^ while excluding the longer “big tau” isoforms predominantly produced outside of the brain, including peripheral nerves and neuronal components that project outside of the brain.^35–37^ In fact, big tau makes up less than 1% of the total brain tau signal but accounts for the vast majority of non-CNS tau.^38^ Additionally, big tau isoforms account for a larger proportion of the total amount of tau protein in the adult human spinal cord than the brain, providing a potential opportunity for differential diagnosis of disorders of the brain vs. spinal cord.^38^

We hypothesized that baseline and longitudinal serum BD-tau levels will be distinctively sensitive to the diverging localization, clinical classification and outcome of acute TBI and SCI. Here, we show that serum BD-tau concentrations accurately enable the differential diagnosis of TBI from SCI among patients admitted in a neuro-intensive care unit (ICU), with temporal dynamics over a six-day period robustly predictive of clinical severity and long-term functional outcomes specifically in TBI but not in SCI. Importantly, serum BD-tau outperformed cerebrospinal fluid (CSF) BD-tau as well as serum NfL, GFAP and UCHL1 as a differential biomarker for acute neurotrauma. Together, serum BD-tau uniquely captures biochemical signatures specific to CNS injury reflected in TBI but not in SCI. Thus, serum BD-tau is a potential precision diagnostic tool to improve accessible neurotrauma care through accurate and rapid stratification of injury types.

## Results

### Cohort characteristics

This study included 364 total participants, with 342 neurotrauma patients admitted to the emergency department (ED) and/or intensive care unit (ICU) of the University of Pittsburgh Medical Center (UPMC) Trauma Center, Pittsburgh, Pennsylvania, USA (n=285 TBI, n=57 SCI), and 22 healthy controls. The TBI group included 12 mild, 6 moderate and 267 severe cases classified using the Glasgow Coma Scale (GCS) criteria (mild: 13-15, moderate: 9-12, severe: 3-8), while the SCI group included 22 AISA-A, 7 AISA-B, 16 AISA-C and 4 AISA-D cases respectively based on the ASIA Impairment Scale (AIS) criteria, with AIS grade missing for 8 SCI cases (see **Methods**).^39,40^ The SCI subtypes C and D were grouped together for sample size purposes. Baseline serum samples were available for all 364 participants, CSF samples for 261 participants, and paired serum-CSF samples for 258 participants, with a mean serum-CSF collection time differences of 3 hours and a maximum of 12 hours. These are hereafter referred to as the “serum cohort,” the “CSF cohort” and the “matched serum-CSF cohort,” respectively, with their demographic characteristics shown in **Extended Data Tables 1-3.** Longitudinal sampling was performed up to 144h post-injury in the TBI and SCI groups, as sample availability permitted.

Age and sex distributions differed across injury groups in the serum cohort, with individuals with SCI being older than both controls and TBI patients (p=0.003), and a higher proportion of females in the controls (p=0.014) (**Extended Data Table 1**). Similar age differences were observed in the CSF cohort (p=0.001) with SCI participants being older than those with TBI, while sex did not differ (p=0.40) (**Extended Data Table 2**). In the matched serum-CSF cohort, SCI participants remained older than TBI participants (p=0.004), but sex was comparable (p=0.30) (**Extended Data Table 3**). Accordingly, age and sex were included as covariates in subsequent analyses.

### Serum BD-tau robustly differentiates TBI from SCI and controls

Baseline serum BD-tau concentrations were significantly elevated in patients with TBI compared with those with SCI or controls (all pairwise comparisons p<0.001; **Fig. 1a**). Within the TBI group, the concentrations showed a stepwise increase with injury severity, with significantly higher levels observed in moderate and severe cases compared with mild cases (**Fig. 1b**). In contrast, serum BD-tau levels in SCI remained low and did not vary across SCI severity (ASIA) subgroups (**Fig. 1b**).

**Fig. 1.**
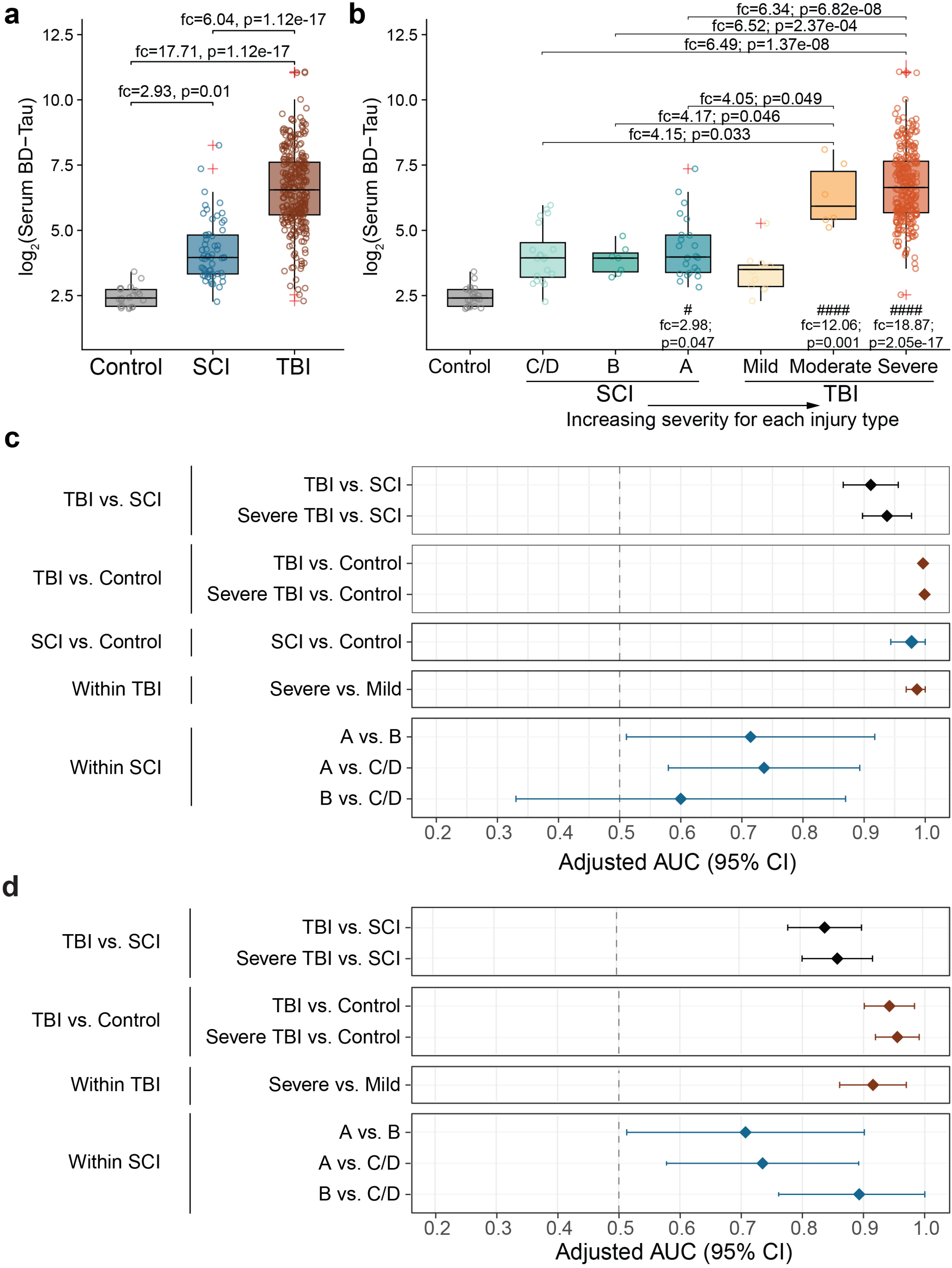
Serum BD-tau levels across control, SCI, and TBI groups. **a-b**, Boxplots show the distribution of log_2_-transformed serum BD-tau concentrations by cohort (**a**) and injury severity (**b**). Boxes represent the interquartile range, and the center lines indicate medians. Statistical comparisons were performed using the Kruskal-Wallis’s test followed by Dunn’s post-hoc tests with false discovery rate (FDR) adjustment, applied to BD-tau values on the original scale. Fold change (fc) denotes the ratio of median values for pairwise comparisons. Asterisks indicate statistical significance (*p < 0.05, **p < 0.01, ***p < 0.001, ****p < 0.0001), and hash symbols indicate comparisons relative to the control group. **c-d**, Forest plot sex- and age-adjusted area under the receiver operating characteristic curve (AUC) values and with 95% confidence intervals for commercial BD-tau (**c**) and in-house BD-tau (**d**) -based discrimination between groups. Unless otherwise specified, TBI and SCI refer to the full cohorts.

Given the marked separation in baseline serum BD-tau concentrations across injury groups and TBI severity, we evaluated the discriminative performance of serum BD-tau using receiver operating characteristic (ROC) analyses. Age- and sex-adjusted ROC analyses demonstrated high accuracy of serum BD-tau in differentiating all TBI from all SCI cases (area under the curve [AUC]=0.91, 95% CI 0.87-0.96) and controls (AUC=0.99, 95% CI 0.99-1.00). These accuracies remained unchanged when limiting the TBI group to the severe TBI cases only (**Fig. 1c**).

Furthermore, serum BD-tau accurately differentiated between the severe and mild TBI groups (AUC=0.99, 95% CI 0.97-1.00). However, as a confirmation of serum BD-tau not being a biomarker for SCI, it had suboptimal AUCs to differentiate SCI subtypes (**Fig. 1c**).

**Supplementary Table 1** summarizes sensitivity, specificity, and related performance metrics; it also reports the corresponding estimates from unadjusted ROC models and 5-fold cross-validation, which were consistent with the primary findings.

To confirm that these findings were not assay-dependent, we repeated the ROC analyses in a sub-cohort (n=246; TBI n=177, SCI n=47 and control n=22) with a different BD-tau assay – an in-house method utilizing a separate set of antibodies.^41^ We observed the same pattern of strong discriminative performance in distinguishing TBI from SCI, and differentiating TBI cases according to injury severity but not SCI subtypes (**Fig. 1d**).

### Serum BD-tau outperforms CSF BD-tau for differential diagnosis of TBI versus SCI

To determine whether the observed diagnostic performance of serum BD-tau reflected a general property of BD-tau release, we compared serum and CSF BD-tau concentrations in participants with paired baseline samples. Serum and CSF BD-tau concentrations were modestly correlated (Spearman ρ=0.30, p<0.001; **Fig. 2a**). Stratified analyses indicated that this association was primarily driven by the TBI group (ρ=0.20, p=0.002) and was not significant in the SCI group (ρ=0.12, p=0.51).

**Fig. 2.**
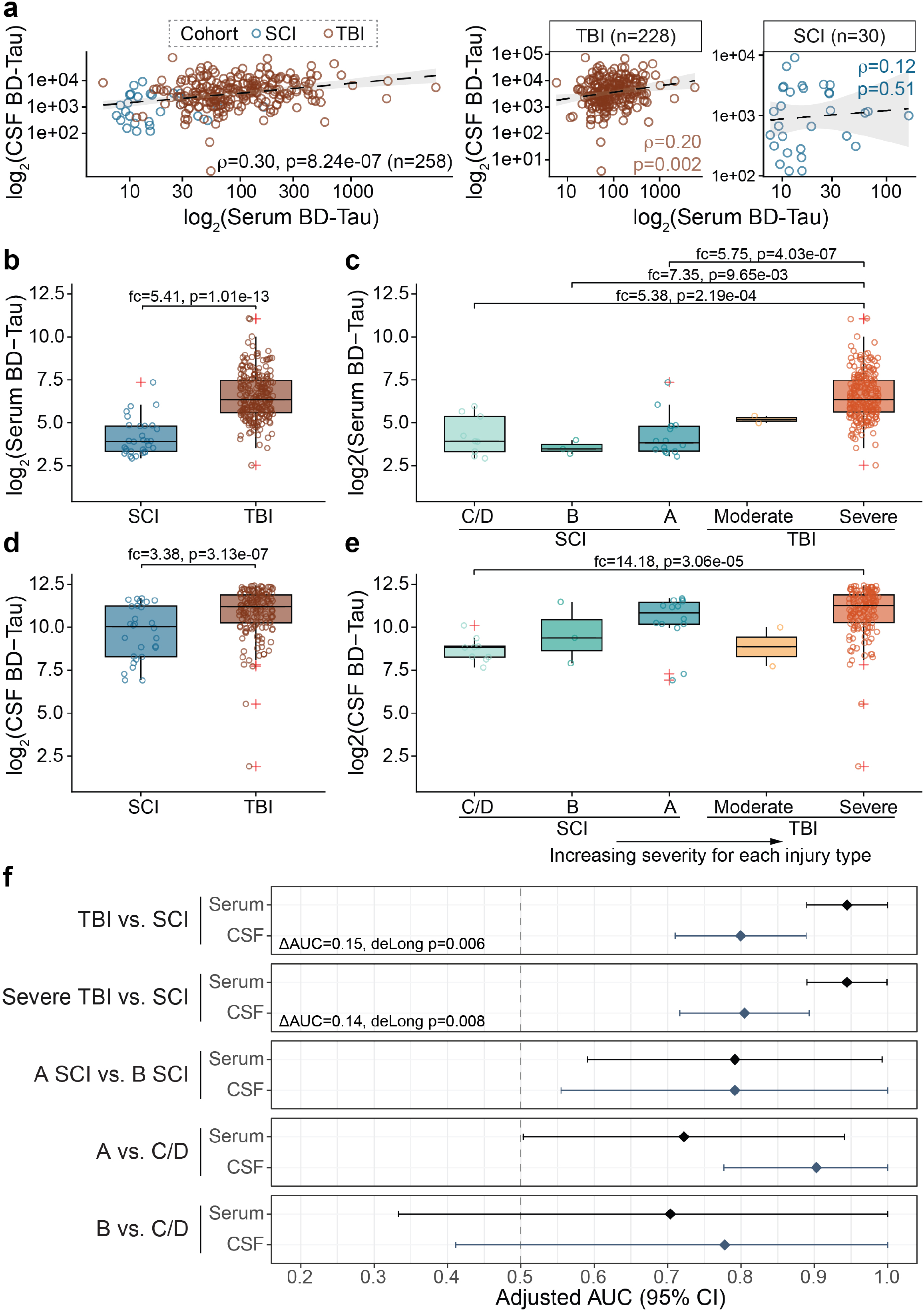
Serum and CSF BD-tau levels measured within 12 hours across control, spinal cord injury (SCI), and traumatic brain injury (TBI) groups. **a**, Spearman rank correlation of log_2_-transformed serum and CSF BD-tau. Spearman’s rho (ρ) values are shown for the overall cohort and stratified by SCI and TBI. Asterisks indicate significance levels: *, p < 0.05; **, p < 0.01; ***, p < 0.001. **c-e**, Boxplots show the distribution of log_2_-transformed serum (**b**-**c**) and cerebrospinal fluid (CSF) (**d**-**e**) BD-tau levels by cohort and injury severity. Boxes represent the IQR, with the center line indicating median. Statistical comparisons were conducted using the Kruskal-Wallis test followed by Dunn’s post hoc tests with FDR adjustment, applied to BD-tau values on the original scale. Fold change (fc) denotes the ratio of median values for pairwise comparisons. Asterisks indicate statistical significance (*p < 0.05, **p < 0.01, ***p < 0.001, ****p < 0.0001). **f**, Forest plots showing sex- and age-adjusted AUC values and with 95% confidence intervals for serum and CSF BD-tau discrimination between groups. Unless otherwise specified, TBI and SCI refer to the full cohorts.

Both serum and CSF BD-tau concentrations were higher in TBI than in SCI (all p<0.001; **Fig. 2b**-**e**). However, the magnitude of separation was substantially greater in serum than in CSF, with larger fold differences between TBI and SCI observed in serum measurements. Consistent with this, ROC analyses showed significantly higher discriminative accuracy for serum BD-tau than for CSF BD-tau in distinguishing TBI from SCI (AUCs = 0.95, 95%CI 0.89-0.99 vs. 0.80, 95%CI 0.71-0.90; **Fig. 2f**).

When stratified by severity, serum BD-tau concentrations were significantly higher in severe TBI than in all SCI subgroups (p<0.001) and did not differ across SCI severity categories (**Fig. 2b**). By contrast, CSF BD-tau concentrations increased with injury severity in both TBI and SCI, with comparable levels observed in severe TBI and the most severe SCI subgroup (ASIA A) (**Fig. 2e**).

### Serum BD-tau outperforms established serum biomarkers including NfL, GFAP and UCHL1

To contextualize this discriminative performance of serum BD-tau, we compared its performance with those of established serum biomarkers of neurotrauma, including neurofilament light (NfL), glial fibrillary acidic protein (GFAP), and ubiquitin C-terminal hydrolase L1 (UCH-L1), in a sub-cohort of 228 participants (TBI: n=162, SCI: n=52, and control: n=14) (see **Supplementary Table 2** for characteristics). In this sub-cohort with paired measures of all four biomarkers, serum BD-tau had a significantly higher discrimination accuracy for TBI versus SCI (AUC=0.82, 95%CI 0.76-0.88) than GFAP (AUC=0.67, 95%CI 0.58-0.76), NfL (AUC=0.53, 95%CI 0.43-0.63, or UCH-L1 (AUC=0.71, 95%CI 0.64-0.79 (**Fig. 3a-b**). Pairwise comparisons using DeLong’s test confirmed the superiority of serum BD-tau over each comparator biomarker (all p<0.01).

**Fig. 3.**
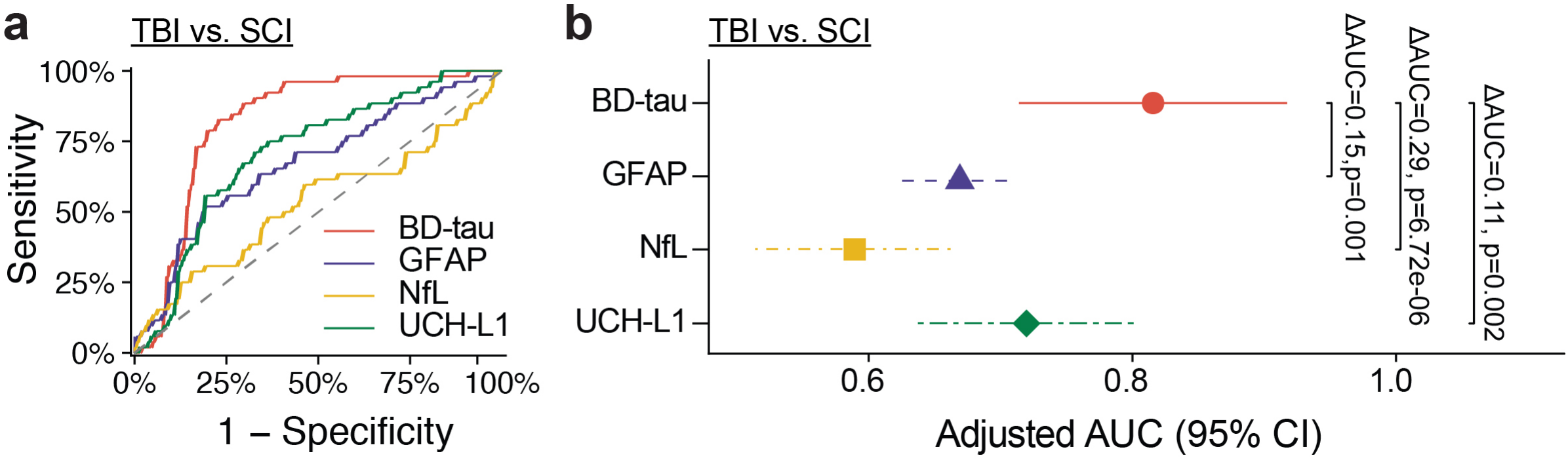
Classification performance of serum BD-tau, GFAP, NfL, and UCH-L1 for separating TBI and SCI. **a**, Receiver operating characteristic (ROC) curves for serum BD-tau, GFAP, NfL, and UCH-L1 discriminating TBI from SCI. **b**, Forest plots show sex- and age-adjusted AUC values and with 95% confidence intervals for serum BD-tau, GFAP, NfL, and UCH-L1 discrimination performance between TBI and SCI. ΔAUC values and asterisks indicate statistically significant differences in AUC between biomarkers based on DeLong’s test for correlated ROC curves (*p < 0.05, **p < 0.01, ***p < 0.001, ****p < 0.0001).

### Temporal serum BD-tau dynamics distinguish TBI from SCI

Consistent with the separation observed at baseline (**Fig. 1**), longitudinal profiling of serum BD-tau up to 144 hours (6 days) post-injury revealed sustained discrimination between injury groups (**Fig. 4a**). Patients with TBI maintained elevated serum BD-tau concentrations across all sampled time points, whereas concentrations in SCI patients remained low and closely approximated those observed in trauma controls. Linear mixed-effects models demonstrated significant group-by-time interactions, indicating distinct temporal trajectories between TBI and SCI (**Fig. 4a**).

**Fig. 4.**
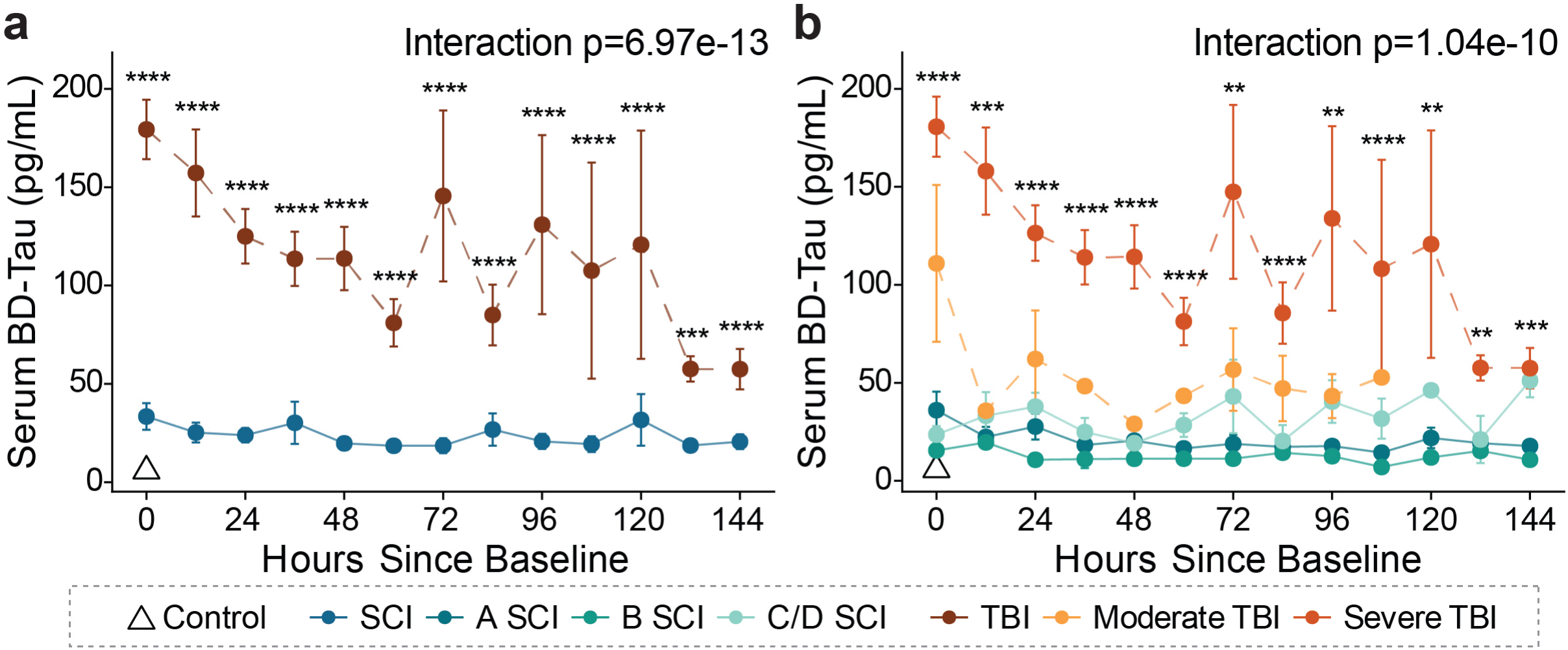
Longitudinal serum BD-tau trajectories by neurotrauma diagnosis. **a-b**, Group-level longitudinal trajectories of serum BD-tau levels measured from baseline through 144 hours (6 days) post-injury by cohort (**a**) and by injury severity, including SCI severity (ASIA A, B, C/D) and TBI severity (moderate and severe), with controls shown for reference (**b**). Points and connecting lines represent group-level mean serum BD-tau values at each time point; error bars indicate ± standard error of the mean. Repeated measurements from individual participants contribute to longitudinal estimates. Inferential analyses were performed on log_2_-transformed serum BD-tau values using linear mixed-effects models with fixed effects for group, time since baseline, and their interaction, and a random intercept for participant. Control participants were excluded from all inferential statistical analyses and are shown for descriptive comparison only. Group-by-time interaction tests for differences in longitudinal BD-tau trajectories between injury groups. Timepoint-specific comparisons were conducted using nonparametric tests on log_2_-transformed values with Benjamini-Hochberg FDR correction applied across time points. Statistical significance is denoted by asterisks (*p < 0.05, **p < 0.01, ***p < 0.001, ****p < 0.0001).

When stratified by severity, moderate and severe TBI showed persistently higher serum BD-tau concentrations throughout the sampling window, with severe TBI exhibiting the highest levels at nearly all time points (**Fig. 4b**). In contrast, serum BD-tau trajectories in SCI were flat across severity subgroups, with no evidence of temporal divergence. Group-specific sample sizes and corresponding statistical results are provided in **Supplementary Table 3**.

### Serum BD-tau trajectories are associated with clinical outcomes after TBI

Given the sustained elevation and heterogeneity of serum BD-tau trajectories among patients with TBI, longitudinal analyses showed distinct serum BD-tau trajectories by 3-month GOS outcome, with linear mixed-effects models identifying a significant outcome-by-time interaction for favorable versus unfavorable recovery (p = 0.022). Patients with unfavorable outcomes generally exhibited higher serum BD-tau concentrations and slower declines over time, although individual timepoint comparisons were not significant after adjustment (**Supplementary Table 4**).

## Discussion

In this study, we have measured serum BD-tau by an immunoassay designed to selectively measure tau protein predominantly originating from the CNS in patients with acute neurotrauma. This is the first reported investigation of BD-tau in SCI and the most comprehensive evaluation in TBI by injury severity, sample time-point and patient number, to date. We have demonstrated that serum BD-tau can reliably differentiate the two common forms of neurotrauma diagnoses in the acute phase, with higher classification accuracy than CSF BD-tau. Notably, serum BD-tau performed significantly superior to serum GFAP, UCH-L1and NfL, the most promising blood biomarkers for neurotrauma currently, in this context of use.

The superior performance of baseline serum BD-tau over GFAP, UCH-L1 and NfL in discriminating TBI from SCI is a novel finding. CSF and blood levels of GFAP, UCH-L1, and NfL post-injury appear to capture shared pathophysiological and mechanistic processes between TBI and SCI.^13,24–26,42^ GFAP and UCH-L1 are released into the bloodstream immediately following neurotrauma in response to astrocytic and neuronal damage. For this reason, they have excellent utility for clinical diagnosis and predicting outcomes across TBI, SCI, and many other acute injury etiologies. Furthermore, NfL, a marker of axonal injury, is released in the acute to sub-acute injury phase, reducing its utility in the immediate hours following TBI and SCI.^23,43^ The ubiquitousness of the temporal signatures of these proteins may limit their specificity for neurotrauma sub-types, and may explain why serum GFAP, NfL and UCHL-1 do not differentiate TBI from SCI as reliably. The distinct serum BD-tau magnitude and temporal trajectory of TBI compared to SCI mirrors the diverging injury pathophysiology, including the mechanisms of injury, the tissue types affected, and thus the proteins released into the bloodstream.

Serum BD-tau more accurately discriminates between severe TBI and SCI compared to CSF BD-tau. We hypothesize that this may be due to the release of this protein biomarker into systemic circulation following blood brain barrier damage after acute injury that circumvents ventricular drainage and proteins of CSF origin.^44^ This is corroborated by the statistically significant correlation between serum and CSF BD-tau in TBI patients, which is consistent with previous reports of BD-tau correlations in paired serum and CSF in chronic neurodegeneration.^45,46^ These findings are similar to our previous results showing that plasma/serum GFAP is a better marker of microglia activation than CSF GFAP.^47^

CSF tau appears relatively understudied, particularly in the severe TBI setting. Of the handful of studies examining the potential of CSF tau as a prognostic indicator, a small cohort showed an association with 12-month GOS-E^48^, while another showed phosphorylated and total tau in CSF slightly outperformed corresponding serum levels for outcome predictions.^27^ Conversely, CSF tau is considered a leading SCI biomarker and is more commonly investigated.^13^ Studies have consistently reported that serum and CSF tau are elevated in TBI compared to controls. In SCI, CSF total-tau distinguished between AIS grades acutely and predicting long-term AIS conversion.^13^ Similarly, our data show CSF BD-tau outperforms serum BD-tau at differentiating ASIA scores. Altogether, BD-tau appears to reflect fundamental differences in injury biology and biomarker kinetics between TBI and SCI.

Measurements of serum BD-tau at 12-hour increments demonstrated differing temporal profiles between moderate/severe TBI (based on initial GCS score) and severe SCI patients. Serum BD-tau levels decreased over time among the TBI patients with the rate of decline dependent on injury severity. Moreover, no TBI group recovered to control levels within the six-day observation period, highlighting the high sensitivity of serum BD-tau to TBI even in less severe presentations. Importantly, while baseline serum BD-tau levels were overlapping for moderate and severe TBI, the longitudinal profiling resolved that individuals with moderate TBI recorded faster serum BD-tau decreases at which point the levels remained high in the severe TBI cases. This approach can be employed clinically to ascertain disease profiles and personalize treatment for TBI patients. Conversely, and remarkably, serum BD-tau levels in SCI remained similar to baseline levels up to 144 hours post-injury and trajectories were similar across ASIA scores or injury location across the spine, indicating that this biomarker is indeed unchanged in SCI.

Serum BD-tau demonstrated distinct prognostic trajectories according to dichotomized long-term outcome classification, defined as unfavorable for GOS 1-3 and favorable for GOS 4-5. While baseline serum BD-tau showed no differences in functional outcome, there was a significant difference in longitudinal trajectory up to 144 hours post-TBI when comparing patients with a favorable versus unfavorable outcome. In a study of 1728 patients across 65 ICUs, fluid biomarker trajectories also had substantial discriminating power above clinical, physiological and imaging variables.^49^ These results support clinical use of serial measurements of serum BD-tau to predict short- and long-term clinical outcomes among TBI patients.

Previous work has primarily focused on establishing prognostic and diagnostic utility of blood-based biomarkers in TBI^50,51^ or SCI alone^52^. By measuring and analyzing TBI and SCI concurrently, our results identify serum BD-tau as an accessible tool to support precision (differential) diagnosis of neurotrauma and to predict outcome among TBI patients. The neurotrauma field aims to modernize characterization of acute TBI with the adaptation of the CBI-M framework.^53^ In addition to the traditional application of clinical (C) and neuroimaging (I) classifications, the CBI-M approach aims to incorporate blood biomarkers (B) and external modifiers (M) to improve clinical management and treatment of TBI.^53^ Our findings provide an opportunity to integrate BD-tau and other blood-based biomarkers into this multi-step workflow for acute neurotrauma care, in conjunction with standard clinical and imaging procedures. For instance, following the generation and external validation of reference values, use of serum BD-tau could establish injury etiology, alongside established markers GFAP and UCH-L1. Together, these findings advance biomarker efficacy in neurotrauma by directly focusing on the unmet clinical need for tools capable of differentially diagnosing these potential co-occurring injury types and improving outcomes following trauma.

This study has several strengths, including a pioneering direct evaluation of TBI and SCI patients in acute intensive care, the inclusion of a relatively large cohort, paired CSF and serum samples, cross-sectional and longitudinal assessments and comparison of serum BD-tau with hallmark blood-based biomarkers of neurotrauma. A few limitations should be noted. There are slight differences in the collection and processing of CSF and blood across the cohorts, primarily that CSF was collected via extra ventricular drain in TBI patients and from lumbar drain in SCI patients, in line with standard of care procedures.^54^ While cell and protein count differ between ventricular and lumbar fluid^55^, tau levels appear to be comparable and highly correlated when directly compared.^56^ Although this study represents the largest and most detailed evaluation of serum BD-tau in acute neurotrauma to date, larger studies are needed to increase statistical power for subgroup analyses and enhance generalizability of findings. Future multicenter studies involving larger and more diverse populations, extended follow-up, and prospective validation will be required to establish the clinical utility of serum BD-tau.

In conclusion, serum BD-tau differentiating TBI from SCI is a novel finding and a significant step toward achieving precision diagnostics in acute neurotrauma patients. Additionally, it can accelerate patient selection for therapeutic clinical trials and observational studies. Finally, these findings demonstrate the potential of circulating biomarkers to improve precision diagnostics in acute neurotrauma care.

## Supporting information

TBI BD-Tau Supplementary Tables

## Data Availability

All de-identified data produced in the present study are available upon reasonable request to the corresponding authors. Access is subject to review and approval in accordance with institutional policies, applicable regulations, and IRB requirements.

## Acknowledgements

TKK and members of the Karikari Laboratory were supported by NIH/NIA (R01 AG083874, U24AG082930, P30 AG066468, RF1AG077474, R01 AG083156, R37 AG023651, R01 AG025516, R01 AG073267, R01 AG075336, R01 AG072641, P01 AG025204), NIH/NINDS (U01 NS131740, U01 NS141777), NIH/NIMH (R01 MH108509), Aging Mind Foundation (DAF2255207), DoD (HT94252320064), the Anbridge Charitable Fund, and a professorial endowment from the Department of Psychiatry, University of Pittsburgh. Control sample collection was funded by Department of Defense (DoD) Award Nos. W81XWH-14-2-0002 (DOO), W81XWH-12-2-0140 (DOO) and W81XWH-18-1-0739 (DOO). SES was supported by NIH T32 HD040686. The content of this article is solely the responsibility of the authors and does not necessarily represent the official views of the funders.

## Author Contributions Statement

Conception, experimental design, and supervision: XZ, DOO, AMP, TKK; Cohort recruitment and retention: AMP, DOO; Plasma sample preparation, biobanking and biomarker data acquisition and processing: SES, XZ, WGB, JDF, MNN, STB, DOO, AMP, TKK; Data compilation, statistical analysis and interpretation of data: RS, SES, XZ; Clinical assessments, coordination and data acquisition: DOO, AMP; Manuscript draft and revision: RS, SES, XZ, WGB, JDF, MNN, STB, DOO, AMP, TKK.

## Methods

### Study Design & Population

The severe TBI cohort samples were obtained from the Brain Trauma Research Center (BTRC) biorepository at the University of Pittsburgh. Patients who were admitted to the level 1 trauma center at the University of Pittsburgh Medical Center Presbyterian hospital and met the following inclusion criteria (1) Initial Glasgow Coma Scale (GCS) score <9 (post-resuscitative; not following commands), (2) Age 16-80, and (3) Positive CT scan for TBI, were prospectively enrolled. Patients were enrolled from 2012-2024 (n=285). Subjects in the moderate TBI group, enrolled in the BTRC study, had an initial GCS score 9-10, but deteriorated to require intracranial monitoring (n=6).

Mild TBI cohort samples were obtained from patients admitted to the University of Pittsburgh Medical Center hospitals with reported trauma or brain injury. Inclusion criteria include 1) Initial Glasgow Coma Scale (GCS) score of 13-15, (2) Age 18 or older, (3) The injury occurred less than 48 hours prior, (4) Loss of consciousness (LOC) and/or post-traumatic amnesia (PTA), (5) Reported mechanism of suspected TBI, and (6) Ability to obtain informed consent. Exclusion criteria include: (1) Significant polytrauma that would interfere with follow-up and outcome assessment, (2) Pregnancy, (3) major debilitating baseline mental health disorders, pre-existing condition or neurological disease impairing baseline awareness, cognition that would interfere with follow-up and the validity of outcome assessment in the opinion of the principal investigator, (4) penetrating TBI, and (5) severe spinal cord injury. Subjects were enrolled from 2018-2024 (n=12).

The SCI cohort samples were obtained from internal SCI biorepository at the University of Pittsburgh Medical Center Presbyterian hospital and met the following inclusion criteria (1) Age 18 and older, and (2) diagnosis of traumatic spinal cord injury (ASIA A, B, C or D) were prospectively enrolled. Subjects were enrolled from 2017-2024 (n=57).

Control subjects blood samples were obtained via 3 parent studies (1) The PRARP (n=9), (2) TEAM TBI (n=5), and (3) ADRC (n=8). These subjects have no history of neurological disorders and no history of TBI hospitalization.

All study protocols were approved by the institutional review board, and written informed consent was obtained from all participants or their legally authorized representatives (IRB: 19030228 (sTBI), 19040200 (mTBI), 19070184, 20020031 (SCI), 19110161, 20050102 (controls)). All consents include language that permits conducting secondary analyses with biofluid samples and de-identified demographic and outcome data. The studies were conducted in accordance with the Declaration of Helsinki.

### Biospecimen Collection and Biomarker Measurement

#### TBI Cohort

All TBI cohort samples were collected and processed by the Neurotrauma Clinical Trials Center (NCTC) at the University of Pittsburgh. Serum specimens (n=3,807) were collected in a 6mL CAT serum clot activator sterile tube (Greiner; cat # 456089) via arterial line or venipuncture. The specimen sat upright for 30-60 minutes and was then centrifuged at 1500 x g for 15 minutes at room temperature. The supernatant was aliquoted into 1.5mL polypropylene tubes in 500uL aliquots and immediately stored at -80°C. CSF specimens (n=6,325) were collected via external ventricular drain and placed in a polypropylene conical tube. The CSF was then aliquoted in 1.5ml polypropylene tubes in aliquots 500uL and immediately stored at -80°C within 30 minutes of collection.

#### SCI Cohort

All SCI cohort samples were collected and processed by the Neurotrauma Clinical Trials Center (NCTC) at the University of Pittsburgh. Serum specimens collected from Spine-Track (n=202) were collected in a 6mL CAT serum clot activator sterile tube via arterial line or venipuncture. The specimen sat upright for 30-60 minutes and was then centrifuged at 1500 x g for 15 minutes at room temperature. The supernatant was aliquoted into 1.5ml polypropylene tubes in 500uL aliquots and immediately stored at -80°C. CSF specimens collected from Spine-Track (n=80) were collected via lumbar ventricular drain. 6mL of CSF was collected in a conical polypropylene tube. Within 30 minutes from collection, the specimen was centrifuged at 1500 x g for 15 minutes at room temperature. The supernatant was then aliquoted into 1.5mL polypropylene tubes in 500uL aliquots and immediately stored at -80°C.

Serum specimens collected from Casper (n=202) were collected via arterial line or venipuncture and sat at room temperature for a minimum of 30 minutes. The sample was then centrifuged at 2,000 x g for 20 minutes at room temperature. The supernatant was then aliquoted into 250uL polypropylene tubes and stored at -80°C. CSF specimens collected from Casper (n=168) were collected by a lumbar intrathecal catheter insertion. 6mL of CSF was collected in a conical polypropylene tube. The CSF was then divided into 1.5mL Eppendorf tubes and then centrifuged at 2,000 x g for 10 minutes. The supernatant was then aliquoted in 200uL increments into 250uL polypropylene tubes and immediately frozen in an ethanol-dry ice bath and stored at -80°C.

### Biomarker Assessments

Serum and CSF samples were brought to room temperature and centrifuged at 10,000× g for 5 min to remove particulates prior to analysis. Serum and CSF BD-tau concentrations were measured on the Quanterix Simoa HD-X Analyzer using the commercially available Quanterix BD-tau Advantage PLUS singleplex assay (lot numbers. 504574 and 504575; Quanterix, Billerica, MA). The same calibrator batch was used across runs. The assay was performed using the recommended 4× dilution for serum and 100× dilution for CSF as previously reported.^57^ For BD-tau singleplex runs, an extended calibration curve beyond the manufacturer-recommended 8-point curve was used, and sample concentrations were calculated from this extended curve.

A subset of patients (TBI: n=162, SCI: n=52, and control: n=14) was additionally analyzed using the Quanterix Neurology 4-Plex D Advantage Plus assay (N4PD+; cat #104822, lot no. 504300) on the Simoa HD-x Analyzer (Quanterix, Billerica, MA). This assay was used to measure serum NfL, GFAP, and UCHL-1 concentrations. Although BD-tau is included in the N4PD+ panel, BD-tau values generated from the multiplex assay were excluded from downstream analyses. For N4PD+ runs, samples were analyzed using the manufacturer-recommended 4× instrument dilution, as previously described.^58^ An extended calibration curve was run, and all N4PD+ samples were refitted to the extended calibration curve.

Each assay plate included calibrators, internal kit controls, and plasma quality-control samples. Samples were loaded onto 96-well plates and assayed in singlicate, whereas calibrators and quality-control samples were assayed in duplicate. Quality-control performance was monitored across runs using measured QC concentrations, intra-plate coefficients of variation, and inter-plate coefficients of variation. Recovery was calculated for quality-control samples but not for calibrators.

For the in-house Pitt-BD-tau assay, after development and validation^59^, BD-tau monoclonal antibody was coupled to paramagnetic beads as the capture agent, and biotin-conjugated Tau12 antibody (Biolegend; cat #SIG-39416) was used as the detector antibody. In vitro phosphorylated recombinant full-length tau441 (SignalChem; cat #T08-50FN-50) was used as the assay calibrator. Serum samples were diluted using Quanterix Homebrew Sample diluent B (lot no. 506221), whereas Quanterix p-tau181 sample diluent (lot no. 104101) was used for both calibrator and detector dilutions.

### Outcome Assessment

Functional outcomes of TBI patients at 3 and 6 months after injury were assessed using the Glasgow Outcome Scale (GOS) score^60^ and dichotomized into two mutually exclusive categories: Unfavorable outcome (GOS=1–3, representing vegetative state or severe disability), and favorable outcome (GOS=4–5, representing moderate disability or good recovery). Missing outcome scores were due to loss to follow-up and/or if information was unavailable in the medical records.

### Statistical Analysis

All statistical analyses were conducted in R (version 4.4.0; R Foundation for Statistical Computing, Vienna, Austria). Analyses were performed using participant-level data. Unless otherwise specified, each biological replicate represents one independent participant. For longitudinal analyses, each participant could contribute one measurement per sampled time point.

Continuous variables were summarized as median with interquartile range (IQR: 25^th^-75^th^ percentile). Categorical variables were reported as counts and percentages. All statistical tests were two-sided. Exact p-values are reported for both significant and non-significant comparisons where relevant; adjusted p-values are reported when multiple-comparison correction was applicated. Statistical significance was defined as p<0.05 after adjustment where applicable. Group comparisons for continuous variables were performed using the Kruskal-Wallis rank sum test, with post-hoc comparisons performed using Dunn’s test and Benjamin-Hochberg false discovery rate (FDR) correction when applicable. Categorical variables were compared using Fisher’s exact test or Pearson’s chi-square test, as appropriate. Spearman rank correlation coefficients were calculated on original concentration scale.

#### Diagnostic performance

Receiver operating characteristic (ROC) curves were constructed to assess the diagnostic performance of BD-tau. Primary comparisons included traumatic brain injury (TBI) versus controls, TBI versus spinal cord injury (SCI), and SCI versus controls. Secondary analyses examined discrimination across injury severity groups, including TBI severity (mild, moderate, severe) and SCI severity (ASIA A, B, C/D), with pairwise comparisons and adjusted for sex and age. Area under the ROC curve (AUCs) with 95% confidence intervals (CIs) was estimated using the DeLong method. Optimal classification thresholds were determined by maximizing the Youden index (sensitivity + specificity - 1). Sensitivity, specificity, and predicted values were estimated at the optimal cutoffs. Pairwise AUC comparisons were performed using the DeLong test for correlated ROC curves.

To assess robustness and mitigate overfitting, ROC analyses were additionally evaluated using resampling approaches. Five-fold cross-validation was performed by partitioning the data into five folds, fitting the model in four folds and evaluating performance in the held-out fold, with performance metrics averaged across folds. In addition, nonparametric bootstrap resampling with 1,000 iterations was used to estimate the variability and confidence intervals of AUC estimates.

#### Longitudinal Trajectory Analysis

Longitudinal BD-tau levels were visualized on the original concentration scale using spaghetti plots with group-level means and standard error (SE) bars at each timepoint. For statistical modeling, BD-tau concentrations were analyzed on the log_2_-transformed scale to reduce right-skewness.

Linear mixed-effects models were fitted with fixed effects for time after injury, group, and the group-by-time interaction, with adjustment for age and sex. Random intercepts and slopes for time were included at participant level to account for within-participant correlation and heterogeneity in individual trajectories. The group-by-time interaction was used to evaluate whether longitudinal BD-tau trajectories differed between groups.

To complement mixed-effects modeling, nonparametric timepoint-specific comparisons were performed to evaluate group differences at individual post-injury time points. At each time point, Kruskal-Wallis tests were used to assess differences in log₂-transformed serum BD-tau concentrations across injury subgroups, followed by FDR correction across time points. Targeted pairwise comparisons between severe TBI and ASIA A SCI were performed at each time point using Wilcoxon rank-sum tests, with FDR adjustment applied across time points.

## Data availability

De-identified data generated in this study can be shared with qualified and identifiable investigators for the purpose of replicating the study results and procedures. Requests should be directed to the corresponding authors, AP and TKK, who will consult the relevant cohort principal investigators as needed. Requests will be reviewed by the investigators and respective institutions to ensure compliance with applicable data-protection regulations, institutional policies, intellectual-property considerations, confidentiality obligations, and the terms of the relevant IRB approvals. These procedures are intended to protect participant anonymity and ensure that data sharing is restricted to approved, secure, and ethnically appropriate use.

## Code availability

No specialized new codes were written specifically for this study.

