## Supplementary material for "Serum brain-derived tau distinguishes acute neurotrauma diagnoses": TBI BD-Tau Supplementary Tables

**Supplementary Table 1**. Discriminative performance of serum BD-tau in ROC analyses

Supplementary Table 1a. Age- and sex- adjusted models

| **Comparison** | **AUC (95%CI)** | **SENS** | **SPEC** | **NPV** | **PPV** | **ACC** | **Cutpoint** |
| --- | --- | --- | --- | --- | --- | --- | --- |
| TBI vs. SCI | 0.91 (0.87-0.96) | 0.85 | 0.9 | 0.51 | 0.98 | 0.86 | 0.84 |
| Severe TBI vs. SCI | 0.94 (0.90-0.98) | 0.89 | 0.9 | 0.6 | 0.98 | 0.89 | 0.83 |
| TBI vs. Control | 1.00 (0.99-1.00) | 0.98 | 1 | 0.82 | 1 | 0.98 | 0.81 |
| Severe TBI vs. Control | 1.00 (1.00-1.00) | 1 | 1 | 0.96 | 1 | 1 | 0.63 |
| SCI vs. Control | 0.98 (0.94-1.00) | 0.98 | 0.96 | 0.96 | 0.98 | 0.97 | 0.61 |
| Severe TBI vs. Mild TBI | 0.99 (0.97-1.00) | 0.99 | 0.92 | 0.79 | 1 | 0.99 | 0.64 |
| A SCI vs. B SCI | 0.71 (0.51-0.92) | 0.46 | 1 | 0.37 | 1 | 0.59 | 0.84 |
| A SCI vs. C/D SCI | 0.74 (0.58-0.89) | 0.59 | 0.9 | 0.67 | 0.87 | 0.74 | 0.61 |
| B SCI vs. C/D SCI | 0.60 (0.33-0.87) | 0.29 | 1 | 0.8 | 1 | 0.82 | 0.48 |

Supplementary Table 1b. Unadjusted models

| **Comparison** | **AUC (95%CI)** | **SENS** | **SPEC** | **NPV** | **PPV** | **ACC** | **Cutpoint** |
| --- | --- | --- | --- | --- | --- | --- | --- |
| TBI vs. SCI | 0.90 (0.85-0.94) | 0.90 | 0.80 | 0.58 | 0.96 | 0.89 | 28.55 |
| Severe TBI vs. SCI | 0.92 (0.88-0.97) | 0.94 | 0.80 | 0.70 | 0.96 | 0.92 | 28.55 |
| TBI vs. Control | 0.99 (0.99-1.00) | 0.98 | 1.00 | 0.79 | 1.00 | 0.98 | 10.76 |
| Severe TBI vs. Control | 1.00 (1.00-1.00) | 1.00 | 1.00 | 0.96 | 1.00 | 1.00 | 11.13 |
| SCI vs. Control | 0.97 (0.93-1.00) | 0.96 | 0.91 | 0.91 | 0.96 | 0.94 | 7.38 |
| Severe TBI vs. Mild TBI | 0.98 (0.96-1.00) | 0.99 | 0.92 | 0.79 | 1.00 | 0.99 | 14.80 |
| A SCI vs. B SCI | 0.60 (0.39-0.82) | 0.32 | 1.00 | 0.32 | 1.00 | 0.48 | 27.77 |
| A SCI vs. C/D SCI | 0.57 (0.39-0.75) | 0.46 | 0.75 | 0.56 | 0.67 | 0.60 | 20.22 |
| B SCI vs. C/D SCI | 0.50 (0.28-0.72) | 1.00 | 0.25 | 1.00 | 0.32 | 0.44 | 8.76 |

Table S1a shows age- and sex-adjusted models, and Table S1a shows unadjusted models. Values are reported as AUC (95% CI), SENS, SPEC, NPV, PPV, ACC, and cutoff. Abbreviations: ROC, receiver operating characteristic; AUC, area under the curve; CI, confidence interval; SENS, sensitivity; SPEC, specificity; NPV, negative predictive value; PPV, positive predictive value; ACC, accuracy; TBI, traumatic brain injury; SCI, spinal cord injury; BD-tau, brain-derived tau.

**Supplementary Table 2**. Baseline demographic characteristics and serum biomarker concentrations in the subset cohort across control, spinal cord injury, and traumatic brain injury groups.

| **Characteristic** | **Overall** | **Control** | **SCI*^a^*** | **TBI*^b^*** | **p-value*^c^*** |
| --- | --- | --- | --- | --- | --- |
|  | N = 228 | N = 14 | N = 52 | N = 162 |  |
| **Sex** |  |  |  |  | <0.001 |
| Female | 60 (26.3%) | 10 (71.4%) | 10 (19.2%) | 40 (24.7%) |  |
| Male | 168 (73.7%) | 4 (28.6%) | 42 (80.8%) | 122 (75.3%) |  |
| **Age** | 44.21 ± 19.15 | 49.57 ± 15.61 | 53.21 ± 20.58 | 40.85 ± 17.98 | <0.001 |
| **Serum BD-tau (pg/ml)** | 97.47 ± 174.76 | 6.10 ± 1.93 | 26.59 ± 43.43 | 128.12 ± 197.91 | <0.001 |
| **GFAP (pg/ml)** | 7,623.25 ± 7,728.00 | 94.65 ± 41.30 | 5,395.94 ± 6,987.07 | 8,988.82 ± 7,767.39 | <0.001 |
| **NfL (pg/ml)** | 211.81 ± 346.00 | 10.08 ± 4.93 | 307.52 ± 517.44 | 198.52 ± 278.03 | <0.001 |
| **UCH-L1 (pg/ml)** | 448.39 ± 1,802.74 | 34.98 ± 33.91 | 165.73 ± 168.24 | 574.85 ± 2,125.16 | <0.001 |

Continuous variables are reported as median with intermediate quartiles (Q1, Q3), and categorical variables are summarized as counts and percentages.

*^a^*The TBI group included participants with mild TBI (n=13), moderate TBI (n=3) and severe TBI (n=146). Because of the small sample size, the moderate TBI subgroup was not analyzed separately in the diagnostic accuracy analysis.

*^b^*The SCI group included participants classified as ASIA-A (n=19), ASIA-B (n=7), ASIA-C/D (n=19), or unavailable ASIA classification (n=7).

*^c^*p-values were calculated using the Kruskal-Wallis rank sum test or Wilcoxon rank-sum test for continuous variables, and Pearson’s chi-square or Fisher’s exact test for categorical variables, depends on sample size.

**Supplementary Table 3**. Longitudinal serum BD-tau trajectories across injury cohorts

Supplementary Table 3a. SCI vs TBI only, Control excluded

| **Time**  **(Hours)** | **SCI** | | | **TBI** | | | **p-value** | **p_adj** |
| --- | --- | --- | --- | --- | --- | --- | --- | --- |
|  | **N** | **Mean** | **SE** | **N** | **Mean** | **SE** |  |  |
| 0 | 54 | 33.4 | 6.8 | 323 | 179.4 | 15.1 | 0 | 0 |
| 12 | 14 | 25.2 | 5.0 | 161 | 157.2 | 22.1 | 7.85E-07 | 1.46E-06 |
| 24 | 31 | 23.9 | 3.7 | 186 | 125.0 | 13.9 | 4.02E-12 | 2.61E-11 |
| 36 | 31 | 30.2 | 10.7 | 148 | 113.6 | 13.8 | 1.32E-11 | 5.73E-11 |
| 48 | 26 | 19.7 | 3.3 | 173 | 113.8 | 16.1 | 1.15E-10 | 2.98E-10 |
| 60 | 29 | 18.5 | 2.3 | 146 | 81.0 | 12.0 | 4.93E-11 | 1.60E-10 |
| 72 | 15 | 18.6 | 3.8 | 146 | 145.6 | 43.4 | 1.00E-06 | 1.63E-06 |
| 84 | 34 | 26.8 | 8.2 | 125 | 85.0 | 15.4 | 2.26E-09 | 4.90E-09 |
| 96 | 14 | 20.8 | 3.8 | 123 | 130.9 | 45.5 | 2.76E-05 | 2.99E-05 |
| 108 | 18 | 19.3 | 4.0 | 90 | 107.6 | 54.9 | 1.25E-05 | 1.62E-05 |
| 120 | 19 | 31.7 | 13.1 | 94 | 120.8 | 58.0 | 2.68E-05 | 2.99E-05 |
| 132 | 11 | 18.7 | 3.1 | 56 | 57.6 | 6.4 | 1.22E-04 | 1.22E-04 |
| 144 | 18 | 20.6 | 3.7 | 47 | 57.5 | 10.3 | 6.78E-06 | 9.79E-06 |

Supplementary Table 3b. SCI vs TBI only, Control excluded

| **Time**  **(Hours)** | **Severe TBI** | | | **A SCI** | | | **p-value** | **p_adj** |
| --- | --- | --- | --- | --- | --- | --- | --- | --- |
|  | **N** | **Mean** | **SE** | **N** | **Mean** | **SE** |  |  |
| 0 | 317 | 180.6 | 15.3 | 23 | 36.0 | 9.5 | 2.25E-17 | 2.93E-16 |
| 12 | 160 | 158.0 | 22.2 | 7 | 22.4 | 5.1 | 1.06E-04 | 1.37E-04 |
| 24 | 182 | 126.4 | 14.1 | 13 | 27.7 | 6.6 | 1.12E-09 | 6.43E-09 |
| 36 | 147 | 114.0 | 13.9 | 16 | 18.3 | 3.0 | 1.48E-09 | 6.43E-09 |
| 48 | 172 | 114.2 | 16.2 | 10 | 20.5 | 5.6 | 7.56E-09 | 2.46E-08 |
| 60 | 145 | 81.3 | 12.1 | 15 | 16.6 | 2.1 | 2.16E-08 | 5.62E-08 |
| 72 | 143 | 147.4 | 44.3 | 6 | 18.9 | 4.6 | 2.22E-05 | 3.61E-05 |
| 84 | 123 | 85.6 | 15.7 | 19 | 17.3 | 1.9 | 2.88E-08 | 6.24E-08 |
| 96 | 119 | 133.9 | 47.0 | 6 | 17.8 | 2.9 | 2.66E-04 | 2.88E-04 |
| 108 | 89 | 108.2 | 55.5 | 10 | 14.4 | 1.8 | 1.25E-04 | 1.48E-04 |
| 120 | 94 | 120.8 | 58.0 | 8 | 21.9 | 5.2 | 4.67E-05 | 6.75E-05 |
| 132 | 56 | 57.6 | 6.4 | 5 | 19.2 | 2.4 | 1.88E-03 | 1.88E-03 |
| 144 | 47 | 57.5 | 10.3 | 8 | 17.7 | 1.9 | 7.47E-07 | 1.39E-06 |

Longitudinal serum BD-tau concentrations are shown by time from injury, excluding controls. Table S3a presents SCI versus TBI comparisons, and Table S3b presents ASIA grade A SCI versus severe TBI comparisons. Values shown are N, mean, SE, p-value, and adjusted p-value (p.adj) at each time point. Abbreviations: SCI, spinal cord injury; TBI, traumatic brain injury; AIS, American Spinal Injury Association Impairment Scale; BD-tau, brain-derived tau; SE, standard error.

**Supplementary Table 4**. Longitudinal serum BD-tau trajectories stratified by favorable versus unfavorable 3-month Glasgow Outcome Scale outcome

| **Time (hours)** | **Comparison** | **N_Favorable** | **N_Unfavorable** | **p-value** | **p_adj** |
| --- | --- | --- | --- | --- | --- |
| 0 | Favorable vs Unfavorable | 51 | 123 | 0.166 | 0.539 |
| 12 | Favorable vs Unfavorable | 31 | 64 | 0.477 | 0.867 |
| 24 | Favorable vs Unfavorable | 31 | 71 | 0.111 | 0.481 |
| 36 | Favorable vs Unfavorable | 25 | 58 | 0.901 | 0.936 |
| 48 | Favorable vs Unfavorable | 25 | 69 | 0.102 | 0.481 |
| 60 | Favorable vs Unfavorable | 20 | 53 | 0.936 | 0.936 |
| 72 | Favorable vs Unfavorable | 19 | 57 | 0.017 | 0.221 |
| 84 | Favorable vs Unfavorable | 17 | 51 | 0.755 | 0.936 |
| 96 | Favorable vs Unfavorable | 20 | 41 | 0.534 | 0.867 |
| 108 | Favorable vs Unfavorable | 18 | 33 | 0.425 | 0.867 |
| 120 | Favorable vs Unfavorable | 16 | 34 | 0.499 | 0.867 |
| 132 | Favorable vs Unfavorable | 8 | 18 | 0.760 | 0.936 |
| 144 | Favorable vs Unfavorable | 8 | 20 | 0.899 | 0.936 |
