## Extended for "Serum brain-derived tau distinguishes acute neurotrauma diagnoses"

**Extended Data Table 1. Baseline demographic characteristics and serum BD-tau concentration across healthy control and injury groups.**

| **Variable** | **Overall** | **Control** | **SCI***^a^* | **TBI***^b^* | **p-value***^c^* |
| --- | --- | --- | --- | --- | --- |
|  | n=364 | n=22 | n=57 | n=285 |  |
| **Age (years)** | 41.0 (26.0, 61.0) | 44.0 (30.0, 58.0) | 54.0 (34.0, 68.0) | 38.0 (25.0, 58.0) | 0.003 |
| **Sex (female)** | 88 (24.2%) | 11 (50.0%) | 12 (21.1%) | 65 (22.8%) | 0.014 |
| **Serum BD-tau (pg/mL)** | 61.7 (28.0, 164.7) | 5.3 (4.2, 6.7) | 15.5 (10.1, 28.2) | 93.7 (48.2, 195.5) | <0.001 |

Continuous variables are reported as median with intermediate quartiles (Q1, Q3), and categorical variables are summarized as counts and percentages.

*^a^*The TBI group included participants with mild TBI (n=12), moderate TBI (n=6) and severe TBI (n=267). Because of the small sample size, the moderate TBI subgroup was not analyzed separately in the diagnostic accuracy analysis.

*^b^*The SCI group included participants classified as ASIA-A (n=22), ASIA-B (n=7), ASIA-C/D (n=6), or unavailable ASIA classification (n=8).

*^c^*p-values were calculated using the Kruskal-Wallis rank sum test or Wilcoxon rank-sum test for continuous variables, and Pearson’s chi-square or Fisher’s exact test for categorical variables, depends on sample size.

**Extended Data Table 2. Baseline demographic characteristics and CSF BD-tau concentration across injury groups.**

| **Variable** | **Overall** | **SCI***^a^* | **TBI***^b^* | **p-value***^c^* |
| --- | --- | --- | --- | --- |
|  | n=261 | n=32 | n=229 |  |
| **Age (years)** | 40.0 (26.0, 60.0) | 55.5 (36.0, 67.5) | 39.0 (25.0, 58.0) | 0.001 |
| **Sex (female)** | 57 (21.8%) | 9 (28.1%) | 48 (21.0%) | 0.40 |
| **CSF BD-tau (pg/mL)** | 3,271.7  (1,410.4, 7,878.9) | 1,049.7  (383.2, 2,649.8) | 3,764.1  (1,896.5, 8,360.0) | <0.001 |

Continuous variables are reported as median with intermediate quartiles (Q1, Q3), and categorical variables are summarized as counts and percentages.

*^a^*The TBI group included participants with moderate TBI (n=2) and severe TBI (n=227). Because of the small sample size, the moderate TBI subgroup was not analyzed separately in the diagnostic accuracy analysis.

*^b^*The SCI group included participants with ASIA-A (n=17), ASIA-B (n=3), and ASIA C/D (n=10).

*^c^*p-values were calculated using the Kruskal-Wallis rank sum test or Wilcoxon rank-sum test for continuous variables, and Pearson’s chi-square or Fisher’s exact test for categorical variables, depends on sample size.

**Extended Data Table 3 Baseline demographic characteristics and matched serum-CSF BD-tau concentration across injury groups.**

| **Variable** | **Overall** | **SCI***^a^* | **TBI***^b^* | **p-value***^c^* |
| --- | --- | --- | --- | --- |
|  | n=258 | n=30 | n=228 |  |
| **Age (years)** | 40.0 (26.0, 59.0) | 54.0 (36.0, 65.0) | 38.5 (25.0, 58.0) | 0.003 |
| **Sex (female)** | 59 (22.9%) | 9 (30.0%) | 50 (21.9%) | 0.3 |
| **Serum BD-tau (pg/mL)** | 70.4 (39.9, 164.0) | 15.1 (10.1, 28.2) | 81.8 (48.3, 180.5) | <0.001 |
| **CSF BD-tau (pg/mL)** | 3,259.0  (1,520.1, 7,564.1) | 1,133.3  (313.2, 2,845.9) | 3,832.1  (1,791.2, 8,193.1) | <0.001 |
| **Hours in between** | 1.7 (0.0, 4.1) | 0.0 (0.0, 0.0) | 2.2 (0.4, 4.8) | <0.001 |

Continuous variables are reported as median with intermediate quartiles (Q1, Q3), and categorical variables are summarized as counts and percentages.

*^a^*The TBI group included participants with moderate TBI (n=2) and severe TBI (n=267). Because of the small sample size, the moderate TBI subgroup was not analyzed separately in the diagnostic accuracy analysis.

*^b^*The SCI group included participants classified as ASIA-A (n=16), ASIA-B (n=3), ASIA-C/D (n=9), or unavailable ASIA classification (n=2).

*^c^*p-values were calculated using the Kruskal-Wallis rank sum test or Wilcoxon rank-sum test for continuous variables, and Pearson’s chi-square or Fisher’s exact test for categorical variables, depends on sample size.
